# Clinical and radiographic features of cardiac injury in patients with 2019 novel coronavirus pneumonia

**DOI:** 10.1101/2020.02.24.20027052

**Authors:** Hui Hui, Yingqian Zhang, Xin Yang, Xi Wang, Bingxi He, Li Li, Hongjun Li, Jie Tian, Yundai Chen

## Abstract

**Objective:** To investigate the correlation between clinical characteristics and cardiac injury of COVID-2019 pneumonia.

**Methods:** In this retrospective, single-center study, 41 consecutive corona virus disease 2019 (COVID-2019) patients (including 2 deaths) of COVID-2019 in Beijing Youan Hospital, China Jan 21 to Feb 03, 2020, were involved in this study. The high risk factors of cardiac injury in different COVID-2019 patients were analyzed. Computed tomographic (CT) imaging of epicardial adipose tissue (EAT) has been used to demonstrate the cardiac inflammation of COVID-2019.

**Results:** Of the 41 COVID-2019 patients, 2 (4.88%), 32 (78.05%), 4 (9.75%) and 3 (7.32%) patients were clinically diagnosed as light, mild, severe and critical cases, according to the 6^th^ guidance issued by the National Health Commission of China. 10 (24.4%) patients had underlying complications, such as hypertension, CAD, type 2 diabetes mellites and tumor. The peak value of TnI in critical patients is 40-fold more than normal value. 2 patients in the critical group had the onset of atrial fibrillation, and the peak heart rates reached up to 160 bpm. CT scan showed low EAT density in severe and critical patients.

**Conclusion:** Our results indicated that cardiac injury of COVID-2019 was rare in light and mild patients, while common in severe and critical patients. Therefore, the monitoring of the heart functions of COVID-2019 patients and applying potential interventions for those with abnormal cardiac injury related characteristics, is vital to prevent the fatality.

## Background

Corona virus disease 2019 (COVID-19) has caused more than 76392 infection cases until 24:00 of Feb. 21 (obtained from Chinese National Health Commission). Despite of respiratory symptoms, multiple organic injury including cellular immune deficiency, coagulation activation, myocardia injury, hepatic injury and kidney injury, has been observed [1]. Researchers reported that about 12% COVID-19 patients suffered acute cardiac injury [2]. Patients with cardiac disease have a higher fatality rate. However, the alerting clinical parameters of cardiac injury after COVID-19 infection and the correlation between cardiac injury and the COVID-19 severity remains to be determined. In this retrospective, single-center study, we recruited 41 consecutive cases of COVID-19 patients admitted to Beijing Youan Hospital, China from Jan 21 to Feb 03, 2020. We selected the indicators, including heart rate (HR), Troponin I (TnI) and epicardial adipose tissue (EAT) observed on chest computed tomographic (CT) scan, that is high related to cardiac injury. Tachycardia, TnI elevation and a trend of lower threshold attenuation EAT density in CT scan have been observed in severe and critical patients. Our results suggested that main attentions should be paid on monitoring the high risk factors of arrhythmia and cardiac function, including HR, TnI, and radiographic feature, that is, EAT density, to protect the COVID-19 patients from cardiac injury.

## Methods

### Data collection

We retrospectively collected the medical records, including epidemiology, clinical, complications, laboratory findings, chest CT scans, therapy procedure and outcome data, of 41 confirmed COVID-19 patients. Clinical outcomes were followed up to Feb 17, 2020. All data were checked by two trained physicians. All patients, in this study, were diagnosed as light, mild, severe and critical cases according to the 6^th^ edition guideline issued by China’s National Health Commission. This study is in compliance with the Institutional Review Board of Beijing Youan Hospital, China.

### Chest CT image acquisition and analysis

All CT images of 41 patients were retrospectively collected. The in-plane size of CT image was 512*512 pixels, and the slice thickness was 0.9 mm. We measured EAT in a range between the bifurcation of pulmonary artery and diaphragm according to the lung CT scan. EAT density was recorded in mean attenuation Hounsfield units (HU). A threshold attenuation value of −195 to −45 HU was defined as the fat-containing areas [3]. According to the size of the heart, we manually depicted the area of intra pericardium on each thin CT image slice. Seven to ten parallel and equidistant axial contour of pericardium were selected by trained cardiologists.

### Statistical analysis

We present continuous measurements as mean±SD if they are normally distributed or median (IQR) if they are not. For continuous variables, comparisons between groups were performed using Kruskal-Wallis test according to the data of normal or non-normal distribution and equal variance, respectively. Categorical data were presented as counts (proportions) and were compared using either a χ2 test or Fisher exact test or Kruskal-Wallis test according to the data. Statistical analyses were conducted using SPSS software (version 19; SPSS Inc, Chicago, IL).

## Results

A total of 41 patients were enrolled, including 19 (46.3%) males and 22 (53.6%) females. 2 (4.88%), 32 (78.05%), 4 (9.75%) and 3 (7.32%) patients were diagnosed as light, mild, severe and critical cases, respectively. The median age was 47 years (35.5-64, range between 15-89 years). 18 patients have been discharged, 21 patients are still in hospital and 2 patients died (their ages were 82 and 65). Old adult male patients are more prone to be in severe conditions (P<0.01). The level of lymphocyte percentage of the severe and critical patients was lower than those of light and mild patients. We found that the level of TnI, one of the cardiac biomarkers, is remarkably higher in critical group (P<0.01) than in other groups, indicative of high risk of cardiac injury. The high level of c-reactive protein (CRP) in critical group (P<0.01) indicated that the occurring of acute inflammation. Note that, the renal and hepatic function biomarkers, creatinine (CRE), alanine aminotransferase (AST) and Aspartate aminotransferase (ALT), were statistically significant (P<0.05). Clinical features and laboratory findings were summarized in Table 1. All these data indicated that COVID-19 could cause multiple organic injury.

**Table 1.** Clinical features and laboratory findings of 41 COVID-19 patients.

|  | Light | Mild | Severe | Critical |
| --- | --- | --- | --- | --- |
| Age (year) | 31.00<br>(4.88%) | 24.97(78.05%) | 68.25 (9.75%) | 70.67 (7.32%) |
| **Gender %** |  |  |  |  |
| Female | 2 (4.88%) | 20 (62.5%) | 0 (0%) | 0 (0%) |
| Male | 0 (0%) | 12 (37.5%) | 4 (100%) | 3 (100%) |
| Respiratory symptoms | 1 (50%) | 30 (93.75%) | 4 (100%) | 3 (100%) |
| Pneumonia | 0 | 29 | 4 | 3 |
| **Laboratory findings** |  |  |  |  |
| White blood cell count<br>(10<sup>9</sup>/L, n=31) | 3.07 | 4.15 (3.40-5.56) | 5.85(4.50-12.87) | 9.40(2.63-12.16) |
| Lymphocyte percentage<br>(n=15) | 34.70 | 28 (22.35-37.35) | 16.60 (11.4-16.8) | 12.70 |
| C-reactive protein (mg/L,<br>n=25) | 1.20 | 14.10 (2.73-23.05) | 55.72 (38.90-98.20) | 191.00 (121.90-211.00) |
| Alanine aminotransferase<br>(U/L, n=22) | 22.00 | 31.50 (18.25-46.50) | 44.50 | 46.00 (31.00-285.00) |
| Aspartate aminotransferase<br>(U/L, n=22) | 20.00 | 26.50 (20.00-41.75) | 53.00 | 70.00 (57.00-380.00) |
| Troponin I (ng/mL, n=20) | 0.01 | 0.01 | 0.10 | 0.54 (0.05-5.90) |
| Creatinine (n=18) | 53.00 | 69 (60-71.75) | 78.00 (40.00-116.00) | 129.00 |
| SpO<sub>2</sub> (% , n=20) | -- | 97(95.38-97.4) | 90.5(88.25-91.5) | 64(60-76) |
| **Oxygen support** |  |  |  |  |
| Nasal Cannula | 0 | 2 | 2 | 0 |
| Non-invasive ventilation | 0 | 0 | 2 | 1 |
| Invasive mechanical<br>ventilation | 0 | 0 | 0 | 2 |

### Cardiac-related chronic diseases in COVID-19 patients

Among the 41 patients, 10 patients have underlying complications, as shown in Figure 1. 1 patient have tumor which has been resected. 9 patients have cardiac-related chronic diseases, 1 patient has type 2 diabetes mellites, 3 patients have hypertension, 2 patients have coronary artery disease, 1 patient have both hypertension and type 2 diabetes mellites, and 2 patients have both hypertension and coronary artery disease. No statistical difference of the classification distribution has been found between patients with any kind of specific cardiac-related chronic diseases (P>0.05). Nonetheless, significant difference of the classification distribution has been observed between patients with cardiac-related chronic diseases and patients with no chronic disease (P<0.05). The number of severe and critical cases are significantly larger in the cardiac-related chronic diseases group (P<0.01). COVID-19 patients with cardiac-related chronic diseases could be more prone to be in the severe or critical cases.

**Figure 1.**
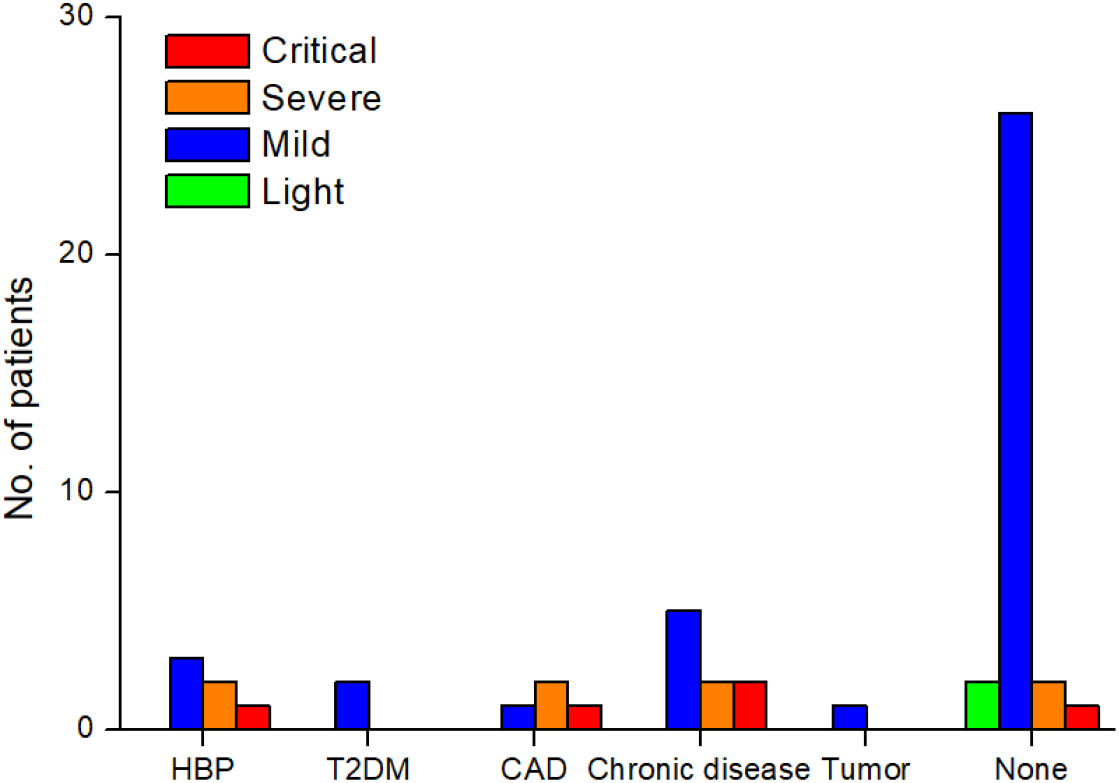
The quantification analysis of cardiac-related chronic diseases in COVID-19 patients.

### Troponin I elevation in severe and critical patients

We collected TnI data from 20 patients. The elevation of TnI elevation was found in 4 patients, containing 1 (1/3) severe patient and 3 (3/3) critical patients, as shown in Figure 2. All 3 critical patients have TnI elevation, and the elevation is statistical significance compared with the light group (p<0.05), the mild group (p<0.01), and the severe group (p<0.01). The TnI elevation in the severe group is not statistical significance compared to the light and mild groups, partly due to the small population of the enrolled patients.

**Figure 2.**
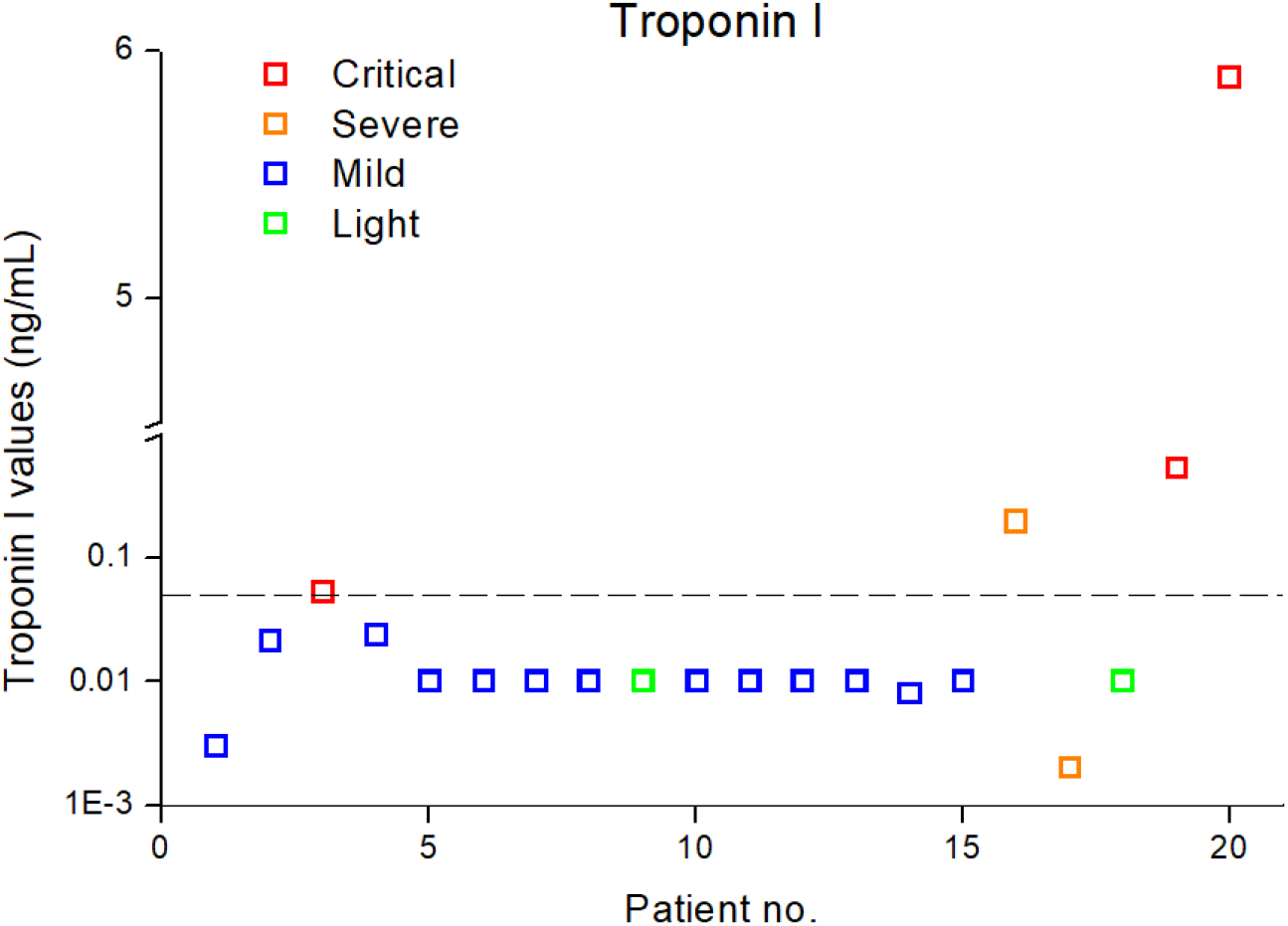
The levels of TnI in COVID-19 patients.

### Tachycardia in severe and critical patients

The heart rates (HR) was documented in 17 patients, and tachycardia was found in 3 patients, of which 1(1/3)in severe group and 2 (2/3) in critical group (Figure 3). The differences of HRs between the critical and the mild (P<0.01) or light group (P<0.01) were statistically significant. The differences of heart rates between the severe and the mild (P<0.01) or light group (P<0.01) were also statistically significant. No significant difference of HR has been found between the severe group and the critical group (P>0.05). As fever itself leads to tachycardia, we analyzed the body temperatures (BT) of different groups. No significant difference of BT has been found between the sever and the mild (P>0.05) or the light(P>0.05) group. There is no significant difference of BT between the mild and the critical group(P>0.05). The rising of HR in COVID-19 patients, which is disproportionate with the increase of the BT. It attracts our attention of tachycardia in the severe and critical COVID-19 patients. Since hypoxia is also a risk factor of elevation of HR, we also analyzed the SpO_2_ in different groups. We found that SpO_2_ is lower in the severe (P<0.01) and critical group(P<0.01), compared with the mild group. Patients in severe and critical groups need more intense oxygen support, and 2 patients in the critical group have invasive mechanical ventilation. However, no significant difference of SpO_2_ has been observed between the severe and the critical group (P>0.05). This is in constant with the change of HR in different groups. More importantly, 2 patients in the critical group had the onset of atrial fibrillation, and the peak heart rates reached up to 123 bpm and 160 bpm respectively. One 82 years old male patient has persistent atrial fibrillation before the infection of COVID-19. Another 65 years old male patient, has not been diagnosed to have atrial fibrillation or other cardiac problems before. Both patients ended up with death. It should be noted that we have to pay great attention on the tachycardia in the severe and critical COVID-19 patients.

**Figure 3.**
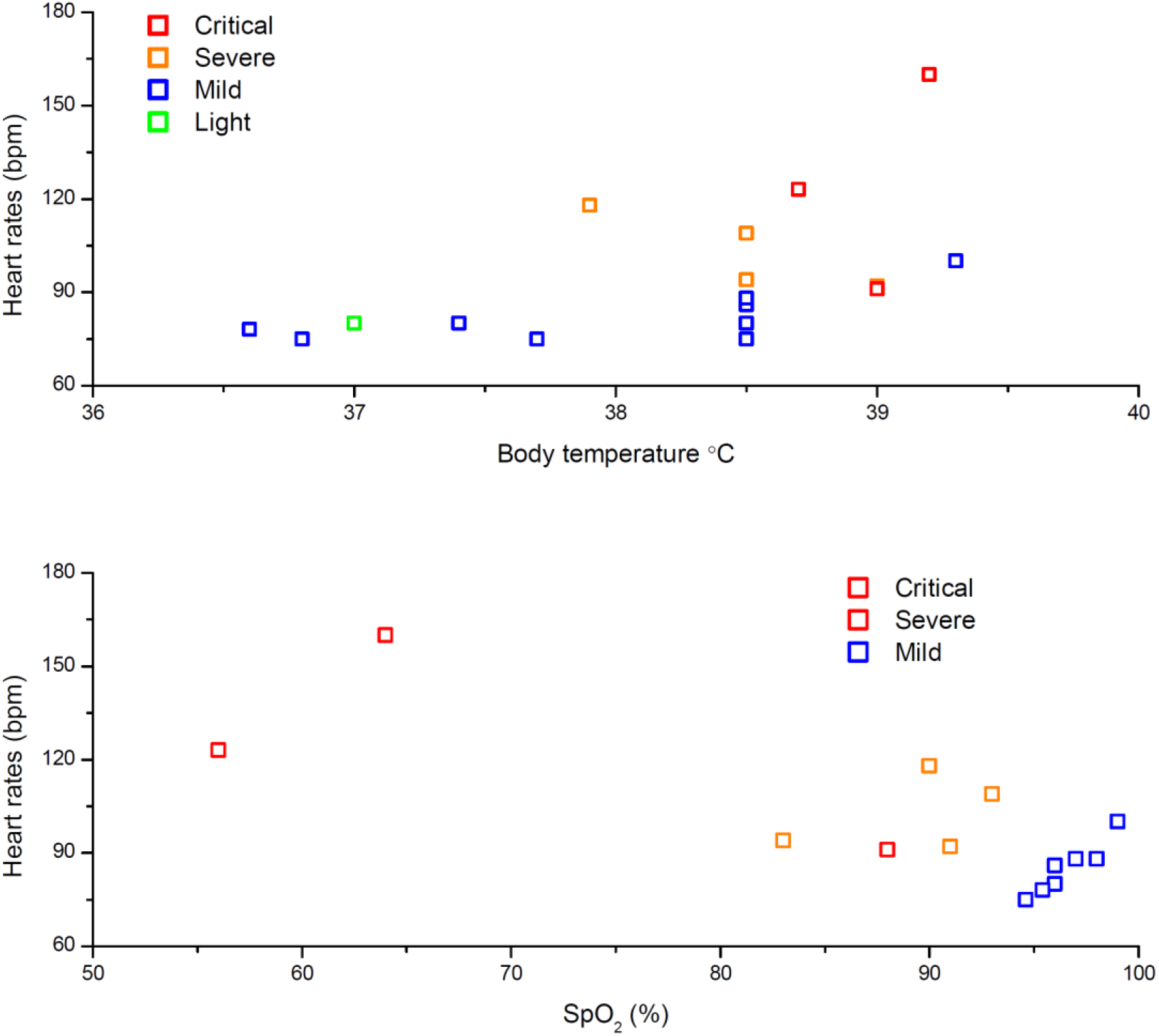
The analysis of heart rates with respect to body temperature (top) and SpO_2_ (bottom) in COVID-19 patients.

### Low EAT density in CT scans for severe and critical patients

We retrospectively collected the plain CT scan results of the hearts in the chest CT scan. In this CT scan example, the CT threshold attenuation value of EAT was measured on the image of the layer range between the bifurcation of pulmonary artery and diaphragm according to the lung CT scan, as shown in Figure 4. We found that the mean EAT density were −98.77 HU and −96.08 HU in the critical (44-year male) and severe (65-year male) groups, respectively (Figure 4c-d), which was lower than patients in the light (−69.37 HU, 44-year female) and mild (−84.76 HU, 37-year male) groups (Figure 4a-b). Patients from severe and critical group had a smaller value than the heathy heart (mean CT value in the range of −70 ∼ −80 HU, p<0.05) [4]. The results indicated that cardiac inflammation may occur in COVID-19 patients.

**Figure 4.**
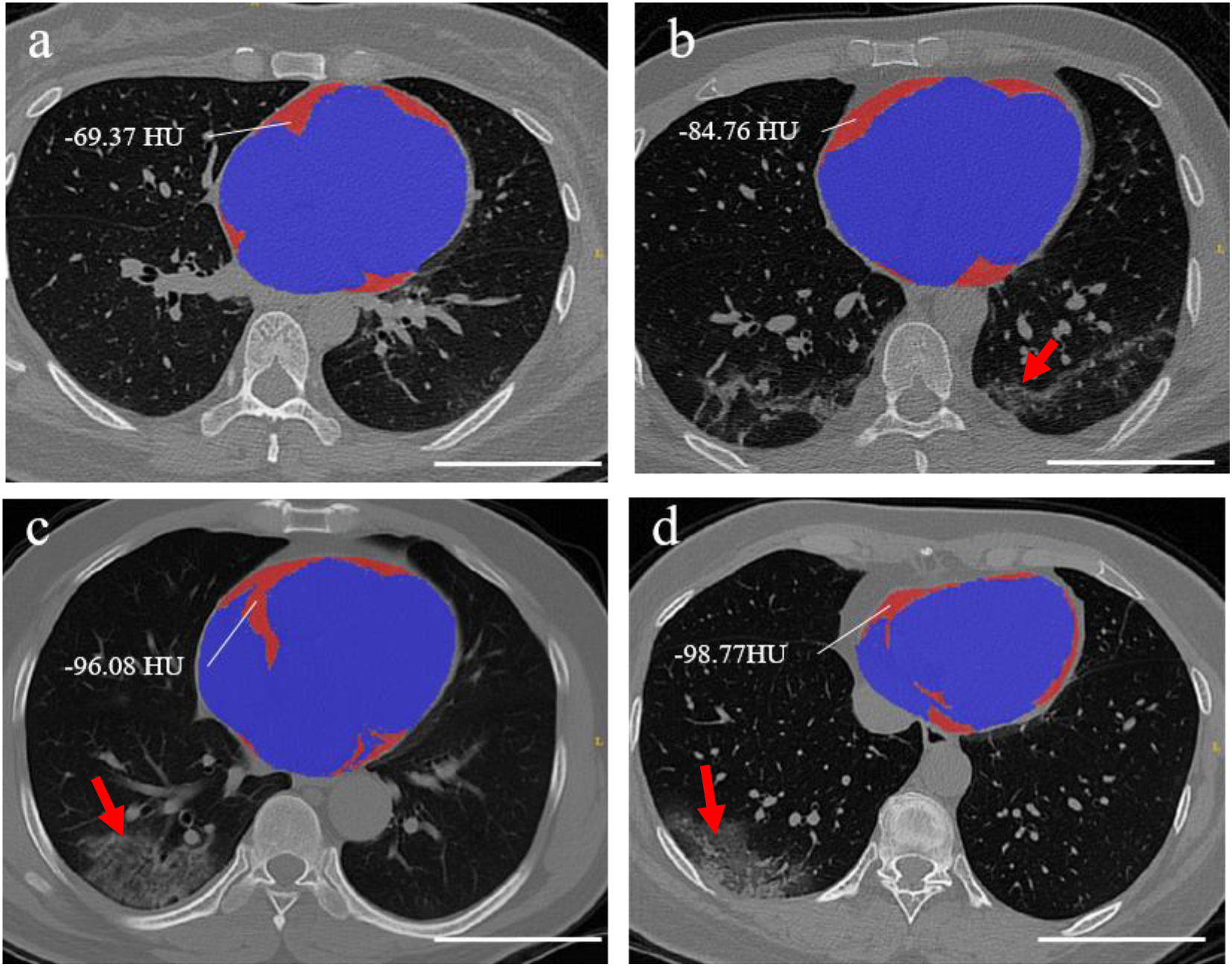
Quantification of EAT density by CT scan. CT scan examples of the patients from light (a), mild (b), severe (c) and critical (d) group of COVID-19 patients. Dark red areas represent the EATs, and blue areas indicate other heart tissues. Red arrows indicate lung lesions, and ground-glass opacity features are obviously presented in severe (c) and critical (d) groups (marked with red arrow). Scale bar, 10 cm.

## Discussion

In this retrospective, single-center study, we investigated the high risk factors, including TnI elevation, tachycardia and lower CT threshold attenuation value of EAT, related to cardiac injury in the COVID-19 patients, especially in the severe and critical groups. Our results suggested the occurrence of cardiac injury in COVID-19 patients. TnI has been widely used as one of the key biomarkers in cardiac injury [2]. In our study, all 3 patients in the critical group and 1 patient in the severe group showed the rising level of TnI. However, patients in light or mild group have no significant TnI elevation. Thus, TnI may be served as severity indicator of cardiac injury in COVID-19 patient. In terms of tachycardia, 2 of 3 patients with Tachycardia had the onset of atrial fibrillation, and their peak heart rates reached up to 160 bpm. However, it is not the case in light and mild group. Compared to another coronavirus, SARS in 2003, tachycardia has also been noticed in patients with SARS [5]. Even in recovery phase the rest HR in SARS patients exceeded 90 bpm, and 72.1% patients have HR above 100 bmp after mild exercises. In our study, the elevation of HR is disproportionate with the increase of the body temperature, however, is related with lower SPO_2_. Higher HR is most likely resulting in worsen prognosis, and tachycardia should be paid extra attention to COVID-19 patients. It is suggested that electrocardiogram (ECG) should also be monitored to specify the classification of tachycardia in the future study.

The analysis of COVID-19 patients with cardiac-related chronic diseases (coronary artery disease, hypertension and type 2 diabetes mellitus) showed that they were more prone to be in the severe or critical conditions. The ALT, AST, and CRE elevation could also be observed in severe and critical group, indicative of multiple organic injury. These findings were consistent with previous studies that COVID-19 caused multiple organic dysfunction and poor prognosis in patients with complications [1]. In a previous study of 1700 SARS patients, the presence of diabetes mellitus and/or cardiac diseases was related both to a higher mortality rate and an adverse outcome [5]. Diabetes mellitus; heart disease; and chronic lung disease were reported more frequently among MERS case-patients than among healthy controls [6, 7]. Patients with cardiac-related chronic diseases should get more intense clinical care in COVID-19 and other coronavirus infected diseases.

Recently, a pathological study of COVID-19 showed that few monocyte cells were found in myocardial interstitial, however, no inflammation cells were observed in myocardial tissue [8]. A previous study found that patients infected with COVID-19 had high amounts of IL1B, IFNγ, IP10, and MCP1. Moreover, patients requiring ICU admission had higher concentrations of GCSF, IP10, MCP1, MIP1A, and TNFα than those non-ICU admission [2]. These previous researches supported that the cytokine storm, not be induced by direct virus infection, but was associated with cardiac inflammation in COVID-19 patients. In addition, the rising level of CRP in COVID-19 patients also indicated the occurrence of inflammation in the circulation. In another aspect, EAT, defined as fat within the pericardial sac, has increased inflammatory cytokine expression in patients with CAD at the tissue level [4, 9]. Thus, to further understand the inflammatory in COVID-19 patients, we quantified EAT density in CT scans, in which the lower CT threshold attenuation value considered as the indicator of inflammation [4]. We found that the mean CT threshold attenuation value of EAT in the critical, severe and mild group, were lower than patients in the light group. Patients from severe and critical group had a lower CT threshold attenuation value than the reported heathy heart. This results suggested that lower EAT density showed the cardiac inflammation in COVID-19 patients.

This study has several limitations. First, only 41 confirmed COVID-19 patients from single center were included. It would be better to include as many patients as possible, even from multicenter, to get a more comprehensive understanding of cardiac injury of COVID-19. Second, more detailed heart function-related data, particularly including ECG or echocardiography, was unavailable at this time. Nonetheless, the data presented in this study allow an early assessment of the clinical characteristics of cardiac injury in COVID-19 patients. Finally, chest CT data of the patients in health was unobtainable to determinate the changes of heart functions with and without COVID-19 infection. In future study, the follow-up chest CT scan during hospitalization could be collected to quantify the change of EAT density in CT scans.

In conclusion, this retrospective, single-center study of 41 confirmed COVID-19 patients has showed the correlation between clinical characteristics and cardiac injury of COVID-19. The high risk factors of cardiac injury including tachycardia, TnI elevation and low EAT density in CT scan, have been observed in the severe and critical COVID-19 patients. Our results suggested that the more attentions shall be applied to protect heart functions, particularly for those COVID-19 patients in severe and critical cases.

## Data Availability

The data that support the findings of this study are available from the corresponding authors upon request. Participant data are without names and identifiers, and they will be made available after approval from the corresponding author and National Health Commission.

## Acknowledgments

We thank all members of the Key Laboratory of Molecular Imaging, CAS for their valuable discussion and comments. We thank all the patients involved in this study.

## References

1. Wang, D., et al., Clinical Characteristics of 138 Hospitalized Patients With 2019 Novel Coronavirus-Infected Pneumonia in Wuhan, China. JAMA, 2020.

2. Huang, C., et al., Clinical features of patients infected with 2019 novel coronavirus in Wuhan, China. Lancet, 2020. 395(10223):p. 497–506.

3. Schlett, C.L. and U. Hoffmann, [Identification and quantification of fat compartments with CT and MRI and their importance]. Radiologe, 2011. 51(5):p. 372–8.

4. Goeller, M., et al., Epicardial adipose tissue density and volume are related to subclinical atherosclerosis, inflammation and major adverse cardiac events in asymptomatic subjects. J Cardiovasc Comput Tomogr, 2018. 12(1):p. 67–73.

5. Chan, J.W.M., et al., Short term outcome and risk factors for adverse clinical outcomes in adults with severe acute respiratory syndrome (SARS). Thorax, 2003. 58(8):p. 686–689.

6. Alraddadi, B.M., et al., Risk Factors for Primary Middle East Respiratory Syndrome Coronavirus Illness in Humans, Saudi Arabia, 2014. Emerging Infectious Diseases, 2016. 22(1):p. 49–55.

7. Hui, D.S., et al., Middle East respiratory syndrome coronavirus: risk factors and determinants of primary, household, and nosocomial transmission. Lancet Infect Dis, 2018. 18(8):p. e217–e227.

8. Xu, Z., et al., Pathological findings of COVID-19 associated with acute respiratory distress syndrome. Lancet Respir Med, 2020.

9. Fitzgibbons, T.P. and M.P. Czech, Epicardial and Perivascular Adipose Tissues and Their Influence on Cardiovascular Disease: Basic Mechanisms and Clinical Associations. Journal of the American Heart Association, 2014. 3(2).

